# A Data-Driven Framework for Identifying Intensive Care Unit Admissions Colonized with Multidrug-Resistant Organisms

**DOI:** 10.1101/2021.09.20.21263595

**Authors:** Çağlar Çağlayan, Sean Barnes, Lisa L. Pineles, Eili Y. Klein, Anthony D. Harris

## Abstract

The rising prevalence of multi-drug resistant organisms (MDROs), such as Methicillin-resistant *Staphylococcus aureus* (MRSA), Vancomycin-resistant *Enterococci* (VRE), and Carbapenem-resistant *Enterobacteriaceae* (CRE), is an increasing concern in healthcare settings. Leveraging electronic healthcare record data, we developed a data-driven framework to predict MRSA, VRE, and CRE colonization upon intensive care unit admission (ICU), and identify the associated socio-demographic and clinical factors using logistic regression (LR), random forest (RF), and XGBoost algorithms. We performed threshold optimization for converting predicted probabilities into binary predictions and identified the cut-off maximizing the sum of sensitivity and specificity. We achieved the following sensitivity and specificity values with the best performing models: 80% and 66% for VRE with LR, 73% and 77% for CRE with XGBoost, 76% and 59% for MRSA with RF, and 82% and 83% for MDRO (i.e., VRE or CRE or MRSA) with RF. Further, we identified several predictors of MDRO colonization, including long-term care facility exposure, current diagnosis of skin/subcutaneous tissue or infectious/parasitic disease, and recent isolation precaution procedures before ICU admission. Our data-driven modeling framework can be used as a clinical decision support tool for timely predictions, identification of high-risk patients, and selective and timely use of infection control measures in ICUs.

## 1. Introduction

The increasing prevalence of multidrug resistant organisms (MDROs), bacteria that are resistant to one or more classes of antibiotics, is an increasingly concerning issue in the community, and in particular, healthcare settings where admitted patients are especially susceptible to developing an infection (Centers for Disease Control and Prevention (CDC) 2013) (Spellberg et al. 2008) (Boucher et al. 2009). These organisms pose a significant threat to patient safety in the form of healthcare-associated infections (Haque et al. 2018), which are associated with considerable morbidity, mortality, and healthcare costs (Klevens, Edwards, and Richards 2007), and have the potential to spread within the community (Molton et al. 2013) (Bassetti, Nicco, and Mikulska 2009).

Two MDROs that are the most prevalent causes of HAIs are Methicillin—resistant *Staphylococcus aureus* (MRSA) and vancomycin—resistant *Enterococcus* (VRE) (Calfee 2012) (Harris et al. 2013), which are currently classified as serious threats by the U.S. Centers for Disease Control and Prevention (CDC) (Centers for Disease Control and Prevention (CDC) 2018). MRSA is reported to cause 80,461 infections and 11,285 deaths per year, and VRE causes 20,000 infections and 11,300 deaths per year (Centers for Disease Control and Prevention (CDC) 2013), with both MDROs being associated with poor treatment outcomes following infections (Cosgrove et al. 2003) (DiazGranados et al. 2005), longer length of hospitalization, and higher healthcare costs (Song et al. 2003) (Cosgrove et al. 2005) (Maragakis, Perencevich, and Cosgrove 2008).

In recent years, Carbapenem-resistant *Enterobacteriaceae* (CRE)—an MDRO class that is highly resistant to carbapenems and other antibiotics reserved for treatment of severe infections—have reached concerning levels in healthcare facilities in the U.S. (Centers for Disease Control and Prevention (CDC) 2015), and around the world (World Health Organization 2017). This trend has prompted the CDC to classify CRE as an urgent threat to public health, its highest risk category (Centers for Disease Control and Prevention (CDC) 2013). CRE is less prevalent than MRSA and VRE, causing 9,000 infections and 600 deaths per year (Centers for Disease Control and Prevention (CDC) 2013), but is an immediate public health threat because infections caused by CRE (e.g., pneumonia, urinary tract infections, bloodstream infections and wound infections (Tischendorf, de Avila, and Safdar 2016)) are very difficult to treat (Jacob et al. 2013) and have been associated with poor treatment outcomes (Morrill et al. 2015) (Borer et al. 2009; Papadimitriou-Olivgeris et al. 2013; Schwaber et al. 2008), and high costs (Bartsch et al. 2017).

Colonized patients carry an MDRO at a detectable level, meaning that a cultured swab sample would test positive but the patient would not show clinical indications of illness caused by an MDRO. Harboring MDROs, these patients are at a risk for subsequent infection, as a significant fraction of MDRO colonization will eventually cause clinically apparent infections that are difficult to treat (Coello et al. 1997) (Diekmann, Heesterbeek, and Britton 2012). They also pose a threat to other patients, as healthcare workers who interact with these patients can become contaminated with the organism and transmit it to other patients. As a result, it is important to rapidly identify and then monitor colonized patients so that the colonization rate is kept under control in hospitals, and the risk of transmission and subsequent infection is minimized (Furuno et al. 2004).

The importation of MDROs into hospitals and other healthcare settings is a major determinant for transmission and outbreak (Tacconelli 2006) (D’agata, Horn, and Webb 2002) (Ziakas et al. 2013). Among hospital departments, intensive care units (ICUs) are the wards where the prevalence of MDROs is particularly high (Zhanel et al. 2008) (Hanberger et al. 2009). Further, patients admitted to ICUs are most vulnerable to develop infections due to these organisms (Vincent et al. 2009; Vincent 2003). Accordingly, ICUs have become a central point of focus for the control and prevention of MDRO colonization and infection within hospitals (Strich and Palmore 2017).

A variety of interventions have been proposed and implemented in order to prevent the transmission of MDROs in ICUs. Effective and commonly utilized interventions include (i) hand hygiene, especially when healthcare workers contact colonized or infected patients (Boyce et al. 2009), (ii) contact precautions (e.g., wearing gloves and gowns) when caring for colonized or infected patients (Siegel, Rhinehart, Jackson, and Chiarello 2007), and (iii) isolation or cohorting of colonized or infected patients (Landelle, Pagani, and Harbarth 2013). Despite their effectiveness, however, these preventive measures are often not applied in a timely manner due to imperfect compliance and the delay (or even failure) to detect patients colonized with an MDRO (Harris et al. 2013).

Surveillance for MDRO colonization is an instrumental practice for detecting patients who may require an intervention (Humphreys 2014) (Pofahl et al. 2009; Schwaber and Carmeli 2013). Yet, the implementation and cost-effectiveness of universal (i.e., active) surveillance strategies, such as screening of all newly admitted ICU patients, has been a controversial topic (Edmond and Wenzel 2013). Some critics argue that the costs associated with universal screening, including the opportunity costs of the human and physical resources being utilized, are likely to outweigh the benefits of active surveillance (Wenzel and Edmond 2010). Accordingly, universal surveillance of all patients may not be feasible to implement in many healthcare facilities due to resource constrains (Tacconelli and Cataldo 2008) (Roth et al. 2016) (Lapointe-Shaw et al. 2017). Instead, targeted surveillance strategies, which offer a cost-effective compromise for detecting asymptomatic colonization, have been advocated by national guidelines (Muto et al. 2003) (Siegel, Rhinehart, Jackson, Chiarello, et al. 2007), (Weber et al. 2007) when a sufficiently accurate method for identifying high-risk individuals is available. Accordingly, rapid identification of patients who are at high risk for MDRO colonization is critical for timely and targeted implementation of screening and other preventive measures, as well as administration of appropriate treatment (e.g., avoiding the misuse of antibiotics).

Given the aforementioned challenges, a system that facilitates timely and reliable identification of newly admitted patients who are likely to be colonized with an MDRO would be quite useful to improve patient safety and effective utilization of critical hospital resources (Delerue et al. 2019). By accurately identifying significant risk factors, this system can help define high-risk subpopulations and hence, could enable the implementation of a cost-effective targeted screening program. Moreover, if highly predictive, it can further be used to immediately initiate clinical interventions, such as contact precautions, as soon as a high-risk individual is admitted to the ICU. Such a real-time system would be particularly useful in ICUs because, currently, identification of colonized patients relies on clinical lab results that usually require at least 1-2 days to process and can delay subsequent action to prevent and control the spread of MDROs.

A particular challenge for the design of a reliable prediction framework is the class imbalance problem that is commonly observed in clinical datasets. Clinical datasets are often not balanced in their class labels, where the predictors and/or prediction outcomes do not make up an equal portion of the data. The imbalance can be particularly large when the prediction outcomes are MDROs, as their prevalence is usually < 15% and can be as low as < 2% as observed in our data. Given that ignoring the class imbalance, especially when it is large, yields poor predictions, it is necessary to consider and address this challenge while developing a prediction framework for accurate and reliable results.

In this study, we developed a data-driven framework to identify patients who are likely to be colonized with VRE, CRE, or MRSA upon ICU admission, leveraging two years of electronic health record (EHR) data from a large academic medical center. Our objective was to develop a modeling framework that can cope with significant class imbalance, commonly observed in clinical datasets, and can be used to (1) to generate timely and accurate predictions for newly admitted ICU patients, and (2) to identify the key socio-demographic and clinical factors affecting the incidence of MDRO colonization. Our framework relied on three supervised machine learning algorithms, specifically regularized logistic regression, random forest, and XGBoost, which were trained on the EHR data and facilitate real-time predictions for newly admitted patients to the ICU.

We achieved the following results for the primary MDRO colonization outcomes: 80% sensitivity and 66% specificity for VRE, 73% and 77% for CRE, 76% and 59% for MRSA, and 82% and 83% for colonization with any MDRO (i.e., VRE, CRE, or MRSA). Moreover, our modeling approach identified long-term care facility exposure, current diagnosis of skin/subcutaneous tissue conditions or infectious/parasitic disease, and recent isolation precaution procedures before ICU admission as key predictors. We were able to detect over 80% of positive MDRO cases upon ICU admission with less than a 20% false-positive rate, which would enable timely and targeted implementation of preventive measures for infection control in ICUs.

The remainder of this article is organized as follows: In Section 2, we present our data and describe our methodology. In particular, in Section 2.1, we introduce our data and describe the clinical and socio-demographic predictors included in our models. Then, in Section 2.2, we introduce the predictive models and then describe the techniques we utilize to improve prediction accuracy and address class imbalance for our application. In Section 3, we present our prediction results and report the key predictors for MDRO colonization in our data set. In Section 4, we summarize our results, discuss the policy implications of our approach and findings, propose directions for future research, and conclude our study.

## 2. Materials and Methods

In this section, we first describe our data source, in Section 2.1, and present the variables and prediction outcomes in our dataset. Then, in Section 2.2, we introduce our modeling framework and describe our methods. In particular, first, we introduce the prediction models we used, and then, discuss our model specification (training) and performance evaluation (testing) stages, describing how we performed hyperparameter tuning, stratified cross-validation, threshold optimization, and finally, out-of-sample evaluations.

### 2.1 Data Description

In this study, we used electronic healthcare record (EHR) data from the University of Maryland Medical Center (UMMC), an academic teaching hospital located in Baltimore, Maryland. Our dataset contained records for 3,958 patients admitted to a surgical or medical ICU in 2017 or 2018. In total, we observed 4,670 individual admissions. Our dataset included the following variables: (1) hospital admission source and type, (2) age, (3) sex, (4) race and ethnicity, (5) region/state of residency, (6) total time admitted during prior ICU and hospital inpatient stays within the previous year, (7) prior antibiotic prescription, (8) diagnoses for prior hospital and/or ICU stays within the previous year, (9) diagnoses for current hospital stay that are present before ICU admission, (10) surgical and medical procedures conducted during prior hospital and/or ICU stays within the previous year, and (11) recent procedures conducted for current hospital stay prior to ICU admission. We treated all predictors utilized in the models as categorical. Descriptive statistics regarding these variables and their categories can be found in Appendix A.

AIn the dataset, all prior and current diagnoses were coded using the International Statistical Classification of Diseases and Related Health Problems (ICD)-10 codification. We used the Agency for Healthcare Research and Quality’s Clinical Classifications Software (CCS) to further categorize the prior and current diagnoses that were present on admission (PoA). The CCS is a diagnosis and procedure categorization catalog (*https://www.hcup-us.ahrq.gov/toolssoftware/ccs10/ccs10.jsp*), mapping the ICD-10 diagnosis codes into 18 categories: (1) Infectious and parasitic diseases, (2) Neoplasms, (3) Endocrine, nutritional, and metabolic diseases and immunity disorders, (4) Diseases of the blood and blood-forming organs, (5) Mental illness, (6) Diseases of the nervous system and sense organs, (7) Diseases of the circulatory system, (8) Diseases of the respiratory system, (9) Diseases of the digestive system, (10) Diseases of the genitourinary system, (11) Complications of pregnancy, childbirth, and the puerperium, (12) Diseases of the skin and subcutaneous tissue, (13) Diseases of the musculoskeletal system and connective tissue, (14) Congenital anomalies, (15) Certain conditions originating in the perinatal period, (16) Injury and poisoning, (17) Symptoms, signs, and ill-defined conditions and factors influencing health status, and (18) Residual or unclassified codes.

We labeled a procedure as recent if it was performed during the current hospital stay. We recorded all recent procedures performed in the hospital inpatient settings prior to the current ICU admission with respect to the ICD-10 Procedure Coding System (PCS), for which each character has a categorical indication. Using the first character of the ICD-10 PCS codes, we classify the recent procedures into eight categories as follows: (i) Medical and Surgical (“0”), (ii) Placement (“2”), (iii) Administration (“3”), (iv) Measurement and Monitoring (“4”), (v) Extracorporeal or Systemic Procedures (“5” and “6”), (vi) Other Procedures (“8”), (vii) Imaging (“B”), and (viii) Other/Miscellaneous (“1”, “7”, “9”, “C”, “D”, “F”, “G”, and “X”). Further, using the first two characters of the ICD-10 PCS codes, we also map the recent procedures into 44 categories (See Appendix A). In our analysis, we include both the single- and double-character based categorizations so that our algorithms can learn which specifications are most important for predicting our MDRO outcomes. We classified prior hospital procedures having the ICD-10 PCS codes in a similar manner as the recent procedures.

Prior outpatient procedures were recorded using the Current Procedural Terminology (CPT) system (*https://www.ama-assn.org/amaone/cpt-current-procedural-terminology*), which we classified into 6 categories: (i) Evaluation and Management, (ii) Anesthesia (iii) Medicine (iv) Radiology (v) Pathology and Laboratory, and (vi) Surgery. The CPT codes for surgery include 18 sub-types, enabling us to construct a more detailed categorization with 23 classes. We used both the 6-class and 23-class CPT codes for our analysis.

The prediction outcomes were colonization with VRE, CRE, or MRSA upon ICU admission, both separately and as an aggregate (union) outcome. Conducting active surveillance in the ICUs, UMMC screened newly admitted patients for colonization with these organisms upon admission and periodically during their stay. At UMMC, active surveillance involves taking routine peri-rectal cultures for VRE and nasal cultures for MRSA on all patients admitted to an ICU at the time of admission, weekly and upon discharge. CRE detection was also primarily done via perirectal swabs and also included clinical cultures (e.g., blood, urine, wound cultures). We identified the positive (i.e., colonized) and negative (i.e., uncolonized) results based on the laboratory tests conducted within two days (i.e., both before and after) of ICU admissions. We limited the time window for the screening results within two days (Paling, Wolkewitz, Bode, et al. 2017; Paling, Wolkewitz, Depuydt, et al. 2017) in an attempt to avoid inclusion of acquisition cases, for which initially susceptible patients acquire an MDRO during their ICU stay. In our data set, 13.03% of patients admitted to the ICU tested positive for VRE, 1.45% for CRE, 7.47% for MRSA, and 17.59% for any MDRO.

### 2.2 Prediction Models, Model Training and Validation, and Threshold Optimization

A variety of techniques have been utilized to analyze complex disease dynamics and quantify its parameters (e.g., the estimation of transmission rate), identify risk factors, and estimate the impact of infection control strategies (van Kleef et al. 2013). These approaches include prediction modeling, computational simulation, and analytic-formula based models such as decision trees (Goodman et al. 2016), artificial neural network (Chang et al. 2011), agent-based simulation for a hospital ward (Barnes, Golden, and Wasil 2010b; Codella et al. 2015) or healthcare system (Lee et al. 2013), dynamic patient and healthcare worker networks (Barnes, Golden, and Wasil 2010a; Cusumano-Towner et al. 2013; Ueno and Masuda 2008), compartmental systems dynamics models (based on ordinary differential equations) (D’Agata et al. 2012; de Cellès et al. 2013), (approximate) Bayesian (computation) techniques (Cooper et al. 2008), and Markov chain based approaches (Kastner and Shachtman 1982; Bootsma et al. 2007). Among these techniques, data-driven prediction models, such as the ones we use in this study, are particularly valuable tools for generating real-time predictions, identifying the significant risk factors, and quantifying their impact on the outcomes under study (Wiens and Shenoy 2017). In addition to these modeling-based approaches, there is also rich clinical literature studying MDRO colonization. See Appendix B for a summary of the clinical studies that assessed the risk factors associated with MDRO colonization, and developed simple clinical prediction rules based on the identified predictors.

We utilized three supervised machine learning (ML) algorithms to predict colonized patients upon ICU admission and to identify significant clinical and socio-demographic factors associated with our outcomes of interest: (1) logistic regression (Reed and Berkson 1929) (Berkson 1944), (2) random forest (Breiman 2001), and (3) XGBoost (Chen and Guestrin 2016). To perform regularization and feature selection for our logistic regression models, we used least absolute shrinkage and selection operator (LASSO), which was originally developed for linear regression (Tibshirani 1996) and then applied to other algorithms including logistic regression (Roth 2004).

For each model, we split the data into an 80% subset for training and cross-validation and a 20% subset for out-of-sample evaluation. We used a 10-fold stratified cross validation scheme for both the tuning of hyperparameters for the algorithm and the optimization of the threshold for classification of the predictions (see Figure 1). We selected the 10-fold due to the relatively small sample size of our data, in an effort to preserve as much data as possible for model training. We selected the stratified scheme to account for the class imbalance in our data, which preserves a similar proportion of the positive outcome for each fold as the complete data set.

**Figure 1.**
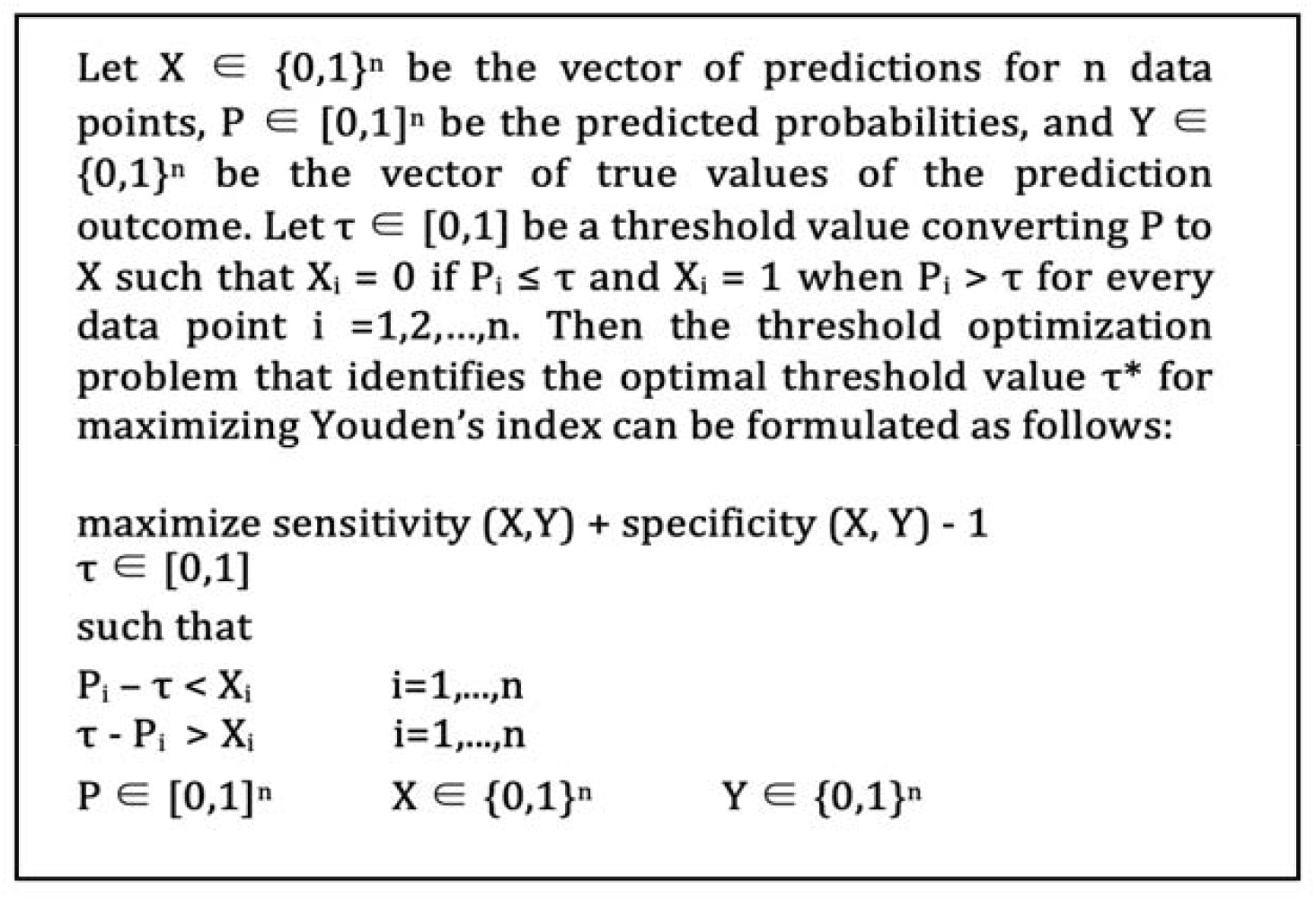
Threshold Optimization Formulation

We defined a grid search for a core set of hyperparameters for each algorithm (see Appendix D), and used the area under the receiver operating characteristic curve (AUC) as the objective function to maximize (out-of-sample) model performance. We selected the hyperparameters achieving the highest mean AUC across the 10 folds for model training.

After choosing the hyperparameters, the next step of the model specification was to identify the ideal cut-off value (i.e., threshold optimization) for converting predicting probabilities into binary predictions. As an initial output, the ML algorithms generates predicted probabilities for the training instances, indicating how likely each patient to be colonized with an MDRO. These predicted probabilities are then translated into binary prediction outcomes using a threshold value. Specifically, observations for which the predicted probabilities are greater than this threshold, τ, are classified as positive (i.e., colonized), otherwise, the patient is assigned to the negative (i.e., susceptible) class. Given the class imbalance observed in our dataset, the default threshold value of 0.5 was unlikely to be effective for our study (see Figure 2). Consequently, we performed an optimization (Sheng and Ling 2006) to search for the best threshold that classifies the predicted probabilities while maximizing the Youden Index (i.e., sensitivity + specificity - 1) for out-of-sample predictions (Youden 1950).

**Figure 2.**
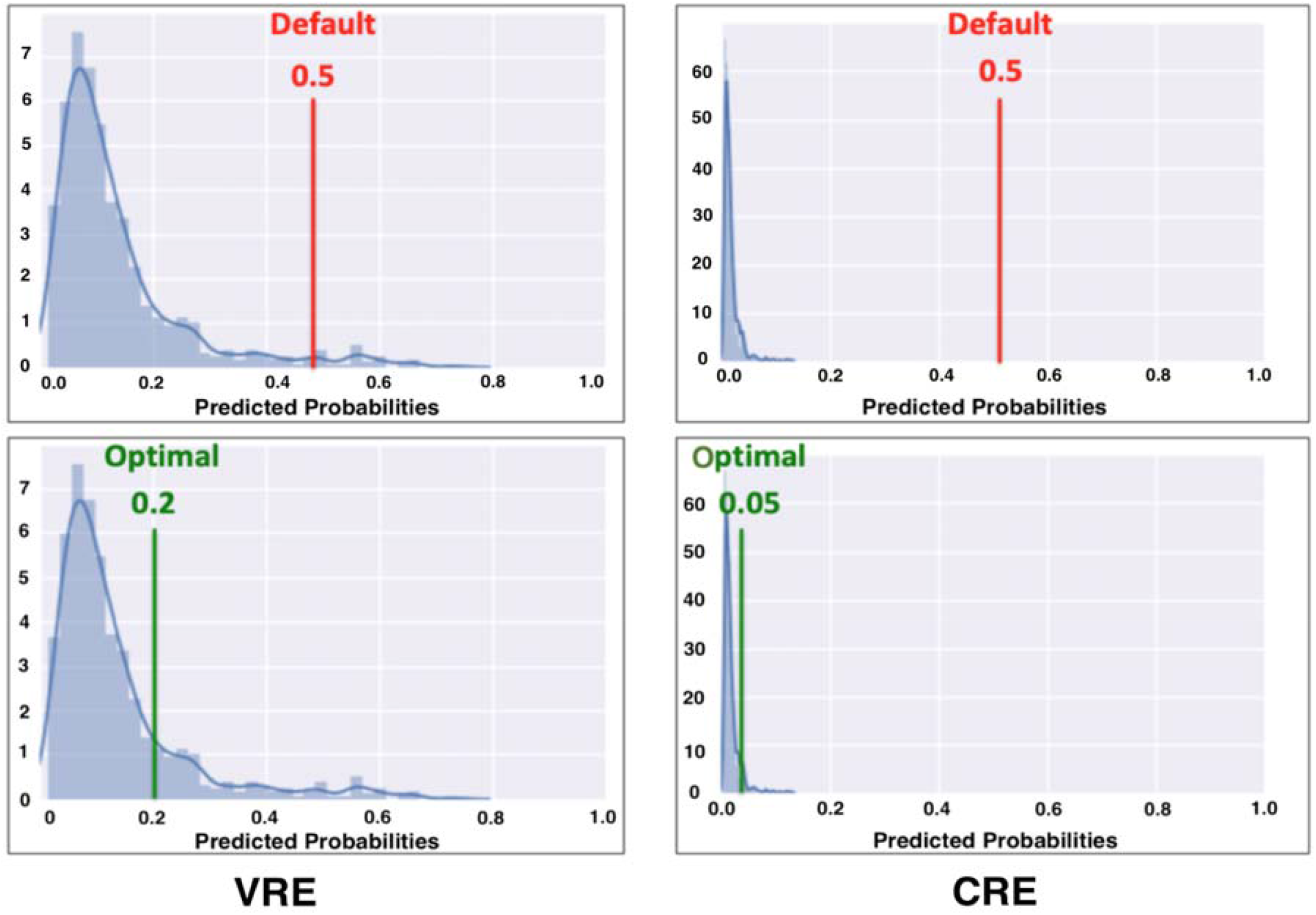
Threshold Value for Converting Predicted Probabilities to Binary Predictions

We performed the threshold optimization out-of-sample using the same 10-fold stratified cross validation scheme used for the hyperparameter tuning. We determined the optimal threshold for each fold using the in-sample predicted probabilities from the 90% subset of training data, and then evaluated the performance (i.e., Youden’s index) of this threshold over the 10% subset of out-of-sample data. We repeated this process for each fold, and selected the mean of these 10 optimal thresholds as our optimized value. We used a bounded numerical search algorithm to solve the optimization problem (Nocedal and Wright 2006), using a lower bound of zero and varying the upper bound for each algorithm to ensure an effective threshold is found. It is noteworthy to emphasize that the upper bound values we considered for each specific outcome were different because the prevalence of the colonized (i.e., positive) instances among VRE, CRE, MRSA, and MDRO were different and directly affected the outcome of the threshold optimization procedure.

Model specification was completed when we determined the hyperparameters, chose the threshold value (for each model), and re-trained the models on the full (80%) training set. Next, we evaluated the performance of the trained models on the (20%) test sets, reporting the AUC, sensitivity, and specificity values obtained. For each MDRO, we conducted a systematic numerical experiment with a range of upper bound values for threshold optimization, and obtained predictions with varying sensitivity and specificity values for VRE, CRE, MRSA, and MDRO (the aggregate prediction outcome). We provide these results in Section 4 for each outcome (e.g., VRE) and algorithm (e..g, XGBoost), and separately, discuss the best performing models for each MDRO.

We also use our modeling framework to identify the key socio-demographic and clinical factors for predicting colonization with VRE, CRE, and MRSA separately and in aggregate. For the LR models, we use odds ratios (ORs), which quantify the associated increase (for values greater than 1) or decrease (for values less than 1) in the likelihood of colonization. For the tree-based models (i.e., RF and XGBoost), we use feature importance (FI), which quantifies the relative frequency that each factor is used to construct the ensemble. Using these two metrics (i.e., OR and FI), we order the identified predictors for each MDRO and report the top five key predictors that are highly ranked across all of the best performing ML models, calculated by the average ranking across the best models.

## 3. Results

We separately predict VRE, CRE, and MRSA colonization upon ICU admission. In addition, combining these three antibiotic-resistant bacteria, we also predict colonization with any of these MDROs (i.e., VRE, CRE, or MRSA) upon ICU admission without specifying the particular organism. As a result, we generate predictions for four cases (namely, VRE, CRE, MRSA, and MDRO) using logistic regression (with LASSO regularization, LR), random forest (RF), and XGBoost algorithms. In Table 1, we summarize the model results for these four outcomes for different upper bound values for the threshold optimization.

**Table 1.**
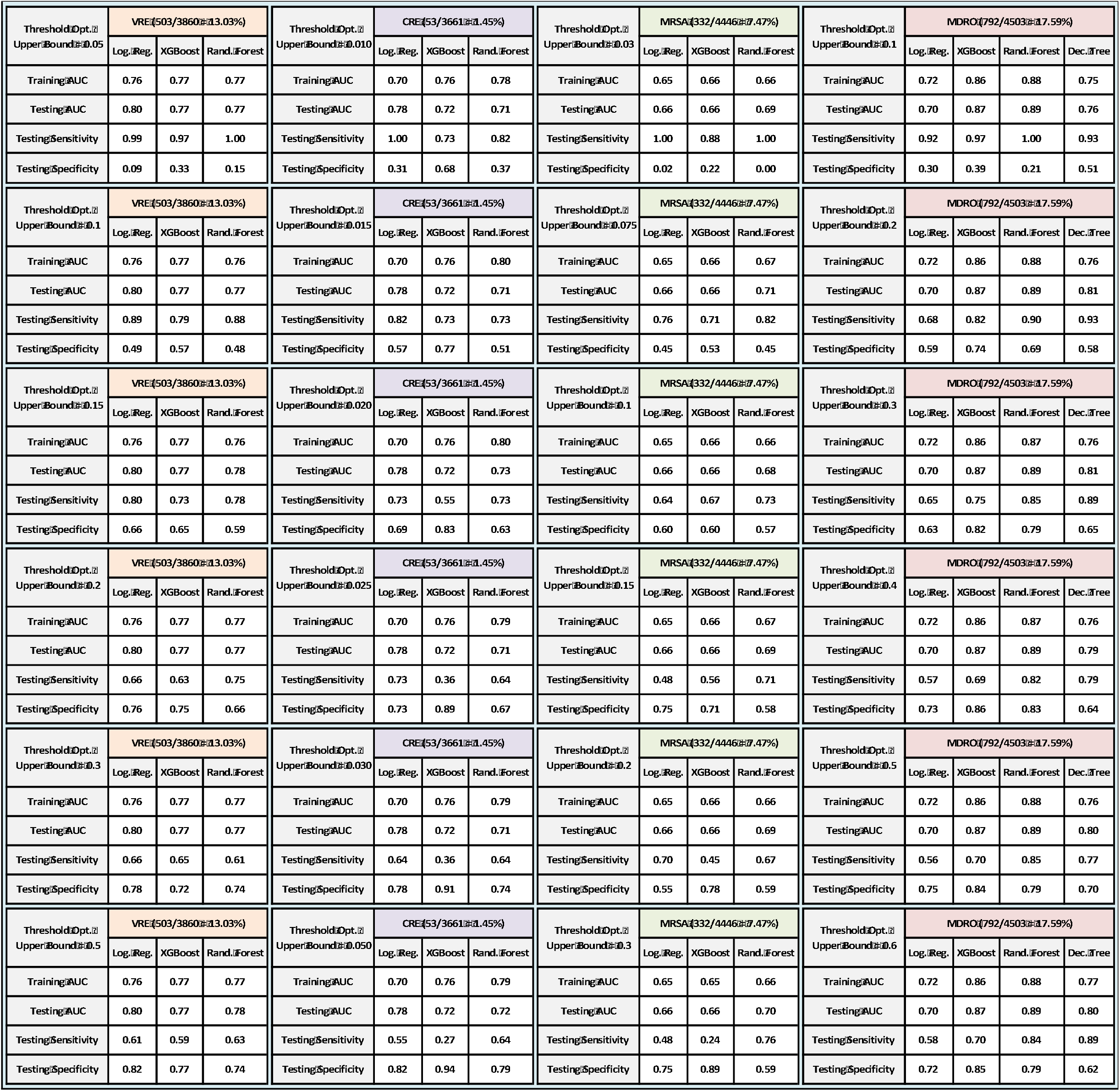
Performance Summary for VRE, CRE, MRSA, and MDRO Colonization **Note:** In each sub-table, we summarize the out-of-sample AUC for the training set, along with the out-of-sample AUC, sensitivity, and specificity for the test set. For each outcome, we include results for different upper bounds for the threshold optimization.

After considering all of the models that we trained for each outcome, we selected the ones with the highest (out-of-sample) Youden index, which we summarize in Table 2. For VRE, the best performing model generated a Youden index of 0.46, achieved via the LR model. By comparison, the RF and XGBoost models generated Youden index values of 0.41 and 0.39, respectively. For CRE, the XGBoost algorithm generate the highest Youden index (0.50), followed by LR (0.45) and RF (0.42). The performance for MRSA is noticeably lower than the other outcomes, for which RF achieved the highest Youden index (0.34). Finally, the prediction models for the aggregate MDRO outcome produced the highest Youden index values when compared to the individual MDRO outcomes, with the RF model (0.65) outperforming the XGBoost (0.57) and LR models (0.30). We note here that the tree-based models performed significantly better than the linear LR model for this aggregated outcome, which is likely due to the former’s natural ability to capture interactions. In an effort to provide support for this hypothesis, we also show the performance of a single classification tree (Breiman et al. 1984) (0.54), which also performs significantly better than the LR model for this particular outcome. On the other hand, for separate VRE, CRE, and MRSA predictions, the single tree models are always dominated by (at least one of) the other algorithms, and hence, not presented in Table 1.

**Table 2.** Performance Summary of the Models with the Highest Youden’s Index

A Data-Driven Framework for Identifying Intensive Care Unit Admissions Colonized with Multidrug-Resistant Organisms
| Models with the Best Youden Index (VRE (503/3860) (13.03%)) |  |  |  |
| --- | --- | --- | --- |
| Best Youden Index | Log. Reg. | XGBoost | Rand. Forest |
| Training AUC | 0.76 | 0.77 | 0.77 |
| Testing AUC | 0.80 | 0.77 | 0.77 |
| Testing Sensitivity | 0.80 | 0.73 | 0.75 |
| Testing Specificity | 0.66 | 0.65 | 0.66 |
| Youden Index | 0.46 | 0.39 | 0.41 |
| Threshold Opt. Bound | 0.15 | 0.15 | 0.20 |

| Models with the Best Youden Index (MRSA (332/4446) (7.47%)) |  |  |  |
| --- | --- | --- | --- |
| Best Youden Index | Log. Reg. | XGBoost | Rand. Forest |
| Training AUC | 0.65 | 0.66 | 0.66 |
| Testing AUC | 0.66 | 0.66 | 0.70 |
| Testing Sensitivity | 0.70 | 0.67 | 0.76 |
| Testing Specificity | 0.55 | 0.60 | 0.59 |
| Youden Index | 0.24 | 0.27 | 0.34 |
| Threshold Opt. Bound | 0.20 | 0.10 | 0.30 |

| Models with the Best Youden Index (CRE (53/3661) (1.45%)) |  |  |  |
| --- | --- | --- | --- |
| Best Youden Index | Log. Reg. | XGBoost | Rand. Forest |
| Training AUC | 0.70 | 0.76 | 0.79 |
| Testing AUC | 0.78 | 0.72 | 0.72 |
| Testing Sensitivity | 0.73 | 0.73 | 0.64 |
| Testing Specificity | 0.73 | 0.77 | 0.79 |
| Youden Index | 0.45 | 0.50 | 0.42 |
| Threshold Opt. Bound | 0.025 | 0.015 | 0.05 |

| Models with the Best Youden Index (MDRO (792/4503) (17.59%)) |  |  |  |  |
| --- | --- | --- | --- | --- |
| Best Youden Index | Log. Reg. | XGBoost | Rand. Forest | Dec. Tree |
| Training AUC | 0.72 | 0.86 | 0.87 | 0.76 |
| Testing AUC | 0.70 | 0.87 | 0.89 | 0.81 |
| Testing Sensitivity | 0.56 | 0.75 | 0.82 | 0.89 |
| Testing Specificity | 0.75 | 0.82 | 0.83 | 0.65 |
| Youden Index | 0.30 | 0.57 | 0.65 | 0.54 |
| Threshold Opt. Bound | 0.50 | 0.30 | 0.40 | 0.30 |

For each model presented in Table 2, the difference between the (out-of-sample) AUC for the (cross-validated) training and testing sets are typically small, suggesting well-trained models. The LR and RF models for CRE demonstrate larger gaps, suggesting that these models might be slightly less robust than others; however, this volatility is likely explained by the extremely low prevalence of positive cases on which to train the models. The best predictions for VRE colonization upon ICU admission are generated by the LR model, which achieves 80% sensitivity and 66% specificity. For CRE, XGBoost produces the best model, having 73% sensitivity and 77% specificity. For MRSA, the RF model performs best, yielding 76% sensitivity and 59% specificity. Finally, the most effective model for the aggregate MDRO outcome is the random forest model, which is capable of detecting 82% of colonized patients with 83% specificity.

In addition to generating predictions, we also use our modeling framework to identify the key predictors for separate and aggregate VRE, CRE, and MRSA colonization. In Table 3, we summarize the top five predictors for the models reported in Table 2, and provide their ranking in the corresponding models as indicated by OR and FI. See Appendix C for the OR and FI values of the factors presented in Table 3.

**Table 3.** Top Five Common Predictors for VRE, CRE, MRSA, and MDRO Colonization **Note:** The “-” sign indicates ranking among the factors that decrease the risk (OR < 1).

| VRE Colonization Upon ICU Admission |  | Relative Ranking |  |  |
| --- | --- | --- | --- | --- |
| Factors | Features | Log. Reg. | XGBoost | Rand. Forest |
| Long-term Care Facility Stay | Yes | 1 | 3 | 1 |
| Recent 1-Digit CD10 Procedure | Other Procedures | 2 | 1 | 2 |
| Current Diagnosis CCS Class | Skin and Subcutaneous Issue | 3 | 2 | 3 |
| Recent 2-Digit CD10 Procedure | Medical/Surgical Anatomical | 6 | 5 | 8 |
| Recent 2-Digit CD10 Procedure | Administration Circulatory | 8 | 4 | 6 |

**Top Five Common Predictors for VRE Colonization Upon ICU Admission**
| CRE Colonization Upon ICU Admission |  | Relative Ranking |  |  |
| --- | --- | --- | --- | --- |
| Factors | Features | Log. Reg. | XGBoost | Rand. Forest |
| Current Diagnosis CCS Class | Skin and Subcutaneous Issue | 2 | 2 | 1 |
| Recent 1-Digit CD10 Procedure | Other Procedures | 3 | 3 | 2 |
| Prior ICU Stay | > 20 Days | 4 | 6 | 5 |
| Long-term Care Facility Stay | Yes | 5 | 6 | 6 |
| Number of Current Diagnosis PoA | > 30 and ≤ 50 | 6 | 8 | 3 |

**Top Five Common Predictors for CRE Colonization Upon ICU Admission**
| MRSA Colonization Upon ICU Admission |  | Relative Ranking |  |  |
| --- | --- | --- | --- | --- |
| Factors | Features | Log. Reg. | XGBoost | Rand. Forest |
| Recent 1-Digit CD10 Procedure | Other Procedures | 1 | 2 | 1 |
| Current Diagnosis CCS Class | Skin and Subcutaneous Issue | 2 | 9 | 2 |
| Current Diagnosis CCS Class | Injury and Poisoning | 7 | 1 | 8 |
| Current Diagnosis CCS Class | Infectious and Parasitic | 9 | 8 | 5 |
| Recent 1-Digit CD10 Procedure | Administration | -3 | 3 | 14 |

**Top Five Common Predictors for MRSA Colonization Upon ICU Admission**
| MDRO Colonization Upon ICU Admission |  | Relative Ranking |  |  |  |
| --- | --- | --- | --- | --- | --- |
| Factors | Features | Log. Reg. | XGBoost | Rand. Forest | Dec. Tree |
| Recent 1-Digit CD10 Procedure | Other Procedures | 7 | 1 | 1 | 2 |
| Current Diagnosis CCS Class | Skin and Subcutaneous Issue | 16 | 2 | 2 | 14 |
| Current Diagnosis CCS Class | Mental Illness | 35 | 6 | 3 | 12 |
| Current Diagnosis CCS Class | Infectious and Parasitic | 57 | 12 | 4 | 16 |
| Sex | Female | 89 | 3 | 5 | 9 |

**Top Five Common Predictors for MDRO Colonization Upon ICU Admission**

Among the recent ICD-10 procedures that were performed during the current hospital stay before ICU admission, procedures categorized as “Other Procedures” in the ICD-10 PCS are among the top five predictors for VRE, CRE, MRSA, and MDRO. In our dataset, a significant proportion of these procedures are “8E0ZXY6”, an ICD-10 code designated for isolation precautions. The patients having a history of a prior colonization or infection for a given MDRO (or are at risk for another indication) are flagged with this code upon admission to the hospital so that they are closely monitored (and if needed, isolated) during their hospital stay. Our results presented in Table 3 show that these patients are at a higher risk for being colonized with an MDRO at ICU admission regardless of the specific indication for which the close monitoring and isolation precautions are put in place.

Another key predictor for VRE, CRE, MRSA, and MDRO colonization is the CCS-based diagnosis category “skin and subcutaneous tissue disease” that is PoA (Table 3). The diagnoses that fall under this CCS category are determined for the current hospital admission and include rash, cellulitis, cutaneous abscess, pressure ulcer, non-pressure chronic ulcer, and other skin conditions. Our finding resonates with the clinical literature and practice as skin and soft tissue infections are amongst the most common bacterial infections, are mostly treated with antibiotics that might cause antimicrobial resistance (Eckmann and Dryden 2010). Further, skin and soft tissue infections are the most frequently reported clinical manifestations of community-acquired MRSA (Fridkin et al. 2005).

For MDRO and in particular MRSA, the CCS-based current diagnosis category “infectious and parasitic diseases” is one of the critical factors that increase the risk of colonization. This category includes diseases such as chronic viral hepatitis C, bacteremia, human immunodeficiency virus (HIV), and sepsis. Patients with these diseases might be at higher risk for MDRO, and in particular MRSA, colonization due to a compromised immune system.

For VRE and CRE, having a prior long-term care facility (LTCF) stay is one of the key predictors for colonization upon ICU admission. This association between VRE or CRE colonization and a previous LTCF stay has been reported by other studies (Prabaker et al. 2012; Tacconelli et al. 2004) (also see Appendix B). High rates of MDRO colonization, debilitating diseases, and the receipt of multiple antibiotics among LTCF residents are likely to be the primary causes of this association both for VRE and CRE colonization (Elizaga, Weinstein, and Hayden 2002).

Other key predictors for VRE were recent procedures “administration circulatory” (ICD-10-PCS ‘30’), such as transfusion, and “medical and surgical anatomical regions, general” (‘0W’), such as drainage, insertion, removal, and transplantation procedures. For CRE, a prior ICU stay longer than 20 days and a total number of diagnoses PoA (i.e., current diagnoses) greater than 30 were two critical factors increasing the risk of colonization. For MRSA, the current diagnosis for “injury and poisoning”, mostly consisting of procedural injuries such as accidental puncture or dural laceration during a procedure, is associated with an increased colonization risk. On the contrary, the recent procedure code for “administration” (i.e., ICD-10 PCS codes with first character “3”) was found to lower the risk of colonization. Finally, female sex and the “mental illness” category for current diagnosis, including diagnosis for cocaine abuse, opioid abuse, poisoning by heroin and psychological disorders, were two other key factors associated with an increased risk for MDRO colonization. Patients in this category (i.e., the “mental illness”) are at higher risk for using injections and causing damage to their skin, which might explain the increased risk for MDRO colonization.

## 4. Discussions, Future Work, and Conclusion

Leveraging a rich EHR data set and supervised ML algorithms, we developed an accurate and interpretable framework for predicting VRE, CRE, and MRSA colonization upon ICU admission. We achieved the following sensitivity and specificity values for VRE, CRE, and MRSA colonization: 80% and 66% for VRE with LR, 73% and 77% for CRE with XGBoost, and 76% and 59% for MRSA with RF. Further, we predicted MDRO (i.e., VRE, CRE, or MRSA) colonization as an aggregate outcome with 82% sensitivity and 83% specificity for MDRO using RF.

Our results indicate that predicting MDRO colonization in aggregate, rather than separately predicting VRE, CRE, and MRSA, achieved the highest prediction accuracy in terms of both AUC and Youden’s index. On the one hand, predicting a specific MDRO would be preferable, as it would enable more customized interventions such as tailored antibiotic therapy. On the other hand, accurately predicting MDRO colonization without specifying whether it is VRE, CRE, or MRSA is still quite important for clinical practice as key interventions for these MDROs are similar such as contact precautions and enhanced environmental cleaning. Accordingly, many infection control measures can be implemented rapidly upon ICU admission for the patients who are suspected to be colonized, and treatment strategies and more advanced interventions can be tailored later as more information becomes available.

In addition to producing timely predictions for newly admitted ICU patients, our ML-based modeling framework can also be utilized to identify the key predictors for VRE, CRE, and MRSA colonization upon ICU admission. We identified several important predictors of MDRO colonization, including long-term care facility exposure, a current diagnosis of skin/subcutaneous tissue or infectious/parasitic disease, and a recent ICD-10 procedure “Other Procedures”, including isolation precaution procedures, as the key predictors for MDRO colonization upon ICU admission. These predictors can help identify ICU patients at high-risk for MDRO colonization and hence, facilitate timely implementation of infection control measures such as selective use of contact precautions, targeted surveillance, and tailored antibiotic therapy.

The primary limitation of our study is that we do not utilize any data on patient medical history outside of UMMC. For example, we do not take into account any antibiotics consumption outside of UMMC or during outpatient visits. Similarly, we do not have information about patients who could have been admitted elsewhere, thus censoring any information about whether they received or underwent additional treatments and procedures in other healthcare facilities. As we utilize administrative data for procedures and diagnoses, which are primarily used for billing, we do not have full access to exact clinical conditions and we do not know the exact reason why a specific procedure was performed or diagnosis was established. Our discussions with clinicians shed some light into these uncertainties but we could not determine the exact details for each individual patient other than what the data conveys. Finally, our data is derived from a single source and we are only able to observe the performance of our modeling framework on an out-of-sample subset from the same facility.

It is noteworthy to emphasize that our study, which focused on predicting MDRO colonization for newly admitted ICU patients, would not prevent the importation of VRE, CRE, and MRSA into the ICU setting. However, by producing reliable predictions and identifying key risk factors for colonization, our approach could enable early detection of colonized patients and facilitate timely and targeted implementation of preventive measures on asymptomatic MDRO carriers. This approach could help reduce transmission of these so-called “superbugs” in ICUs, which we plan to further analyze via a comprehensive simulation model in future efforts, and would particularly be useful for healthcare settings where active surveillance is not performed.

Traditionally, many prediction rules, developed as a decision support tool for clinicians, are designed to be very simple, relying on only a small number of variables, for practicality. Yet, with the increasing availability of electronic healthcare record (EHR) data and the expansion of modern database and software systems, the use of data-driven prediction models and other analytical and computational methods for the identification, control, and prevention of MDROs and other HAIs has been increasing (van Kleef et al. 2013). As a result, a growing number of healthcare facilities are capable of generating more complex prediction models in an automated fashion. Accordingly, taking advantage of the advances in computational and data recording technologies, many healthcare organizations can use our data-driven prediction framework to produce real-time predictions and identify the high-risk patients for MDRO colonization.

There are three research directions that we plan to pursue in near future: First, we will study the acquisition outcomes, where we focus on the ICU patients who were initially colonization-free but acquired VRE, CRE, or MRSA colonization during their ICU stay. Second, we will develop a comprehensive agent-based simulation model to analyze MDRO colonization and infection in ICUs and assess the impact of commonly utilized prevention and control measures on MDRO transmission. Finally, we plan to acquire more data from another major healthcare facility and conduct a similar study by leveraging this additional dataset. This will not only enable us to enlarge the size our dataset, leading to more accurate predictions, but will also give us an opportunity to assess the generalizability of our findings and help us develop more robust predictions.

To summarize, in this study, we proposed a data-centric modeling framework to predict VRE, CRE, and MRSA colonization upon ICU admission and identify the associated risk factors. We achieved the highest prediction accuracy, measured by Youden’s index, when we predicted VRE, CRE, and MRSA colonization combined as an aggregate outcome. Capable of coping with significant class imbalance, a feature commonly observed in clinical datasets, our framework can be used as a clinical decision support tool to provide accurate on-time predictions especially if it is regularly updated and trained off-line as additional (i.e., more recent) data become available. It can further be used to identify the key risk factors and define high-risk populations, for which targeted interventions can be implemented rapidly to reduce transmission of MDROs in ICUs.

## Data Availability

Data cannot be shared publicly because of private ownership. Data were obtained via electronic healthcare record (EHR) data from the University of Maryland Medical Center (UMMC), an academic teaching hospital located in Baltimore, Maryland.

## Acknowledgement

This study is supported by the U.S. Centers for Disease Control and Prevention (CDC) Modeling Infectious Disease (MInD) Network prime award number 1U01CK000536. The content is solely the responsibility of the authors and does not necessarily represent the official views of the CDC.

